# Household food insecurity risk indices for English neighbourhoods: measures to support local policy decisions

**DOI:** 10.1101/2022.04.06.22273530

**Authors:** Dianna M Smith, Lauren Rixson, Grace Grove, Nida Ziauddeen, Ivaylo Vassilev, Ravita Taheem, Paul Roderick, Nisreen A Alwan

## Abstract

**Background:** In England, the responsibility to address food insecurity lies with local government, yet the prevalence of this social inequality is unknown in small subnational areas. In 2018 an index of small-area household food insecurity risk was developed and utilised by public and third sector organisations to target interventions; this measure needed updating to better support decisions in different contexts.

**Methods:** We held interviews with stakeholders (n=11) and completed a scoping review to identify appropriate variables to create an updated risk measure. We then sourced a range of open access secondary data to develop an indices of food insecurity risk in English neighbourhoods. Following a process of data transformation and normalisation, we tested combinations of variables and identified the most appropriate data to reflect household food insecurity risk in urban and rural areas.

**Results:** Eight variables, reflecting both household circumstances and local service availability, were separated into two domains with equal weighting for a new index, the Complex Index, and a subset of these make up the Simple Index. Within the Complex Index the Compositional Domain includes population characteristics while the Structural Domain reflects access to resources. The Compositional Domain is correlated well with free school meal eligibility (r_s_=0.705) and prevalence of childhood obesity (r_s_=0.641). This domain was the preferred measure for use in most areas when shared with stakeholders, and when assessed alongside other configurations of the variables. Areas of highest risk were most often located in the North of England.

**Conclusion:** We recommend the use of the Compositional Domain for all areas, with inclusion of the Structural Domain in rural areas where locational disadvantage makes it more difficult to access services. These measures can aid local policy makers and planners when allocating resources and interventions to support households who may experience food insecurity.

## Introduction

### Food insecurity and health

Hunger and poverty feature prominently in the Sustainable Development Goals, outlining the need to end both experiences globally by 2030(1). Although the focus may traditionally be on Low- and Middle-Income Countries (LMIC), these aims are also relevant for High Income Countries (HIC), such as the UK. UK Stakeholders for Sustainable Development in 2019 pointed to the high levels of household food insecurity in the UK compared to the rest of Europe(2). Here, the term food insecurity reflects the inability of an individual or household to access food of sufficient nutritional quality and quantity using socially acceptable options. Instead, they may need to access high interest loans, or food aid such as food banks – or go hungry(3). The term is sometimes used interchangeably with food poverty to reflect the perspective that people who are experiencing food insecurity often do so because of economic constraints, and it is desirable to include ‘poverty’ in the term to capture this lack of income. Current data place UK household food insecurity prevalence at 8% of households pre-pandemic (4) and up to 9.7% during the pandemic (5).

Food insecurity is a problem that is not easily addressed due to many contributing factors – income, location, personal circumstances which put pressure on resources – and the anticipated solutions which are largely focused on supporting personal income. Conceptually, the causes of and solutions to food insecurity in households have predominantly focused on the cost and availability of food, drawing on ideas of food deserts and food ladders. Food deserts have a long history in the UK with contested definitions around spatial access to affordable, healthy food (see (6)). In contrast, possible solutions to household (and community) food insecurity are described as ladders, with rungs to support households through direct and indirect interventions ranging from free at point of use food banks to development of community food systems (7).

While healthcare is provided by the state in the UK, and largely free at the point of use, food is not. The aim is for people who struggle to afford necessities such as food and housing to use the welfare system to increase their income to a level that enables them to at least meet basic needs. However, there is no requirement for the state to provide food in line with healthcare and housing (noting limited supply of state housing provision (8)). This has created a debate about the Right to Food and whether it should be enshrined in law across all nations of the UK (9). Arguably, this most basic need should be equally protected as it is central to a person’s health and wellbeing. Although food is not within the remit of the government’s required provision such as housing and education, third sector responses to food insecurity such as food banks, the Holiday Activity Fund and voucher schemes during the 2020 lockdown, community fridges and food pantries are often supported in part by local governments as part of their public health strategy. This is in the context of reduced income to local authorities, and the impact of funding cuts which are geographically uneven (10).

What is absent for local governments is the data on household food insecurity *within* their local areas. This would enable allocation of increasingly stretched resources to the areas or populations where food insecurity is greatest and was the motivation behind an earlier food insecurity risk measure (11). A place-focused measure was initially called for in 2013 as part of the terms of reference for the All Party Parliamentary Group on Hunger and Food Poverty (12). Such data are necessary as, following the devolution of public health resources to local governments with the 2012 Health and Social Care Act, the task to prioritise funding on the basis of population needs was shifted to local authorities (13). Crucial to local decision making is reliable data or estimates of health outcomes or related social inequalities. The need for small area data is not limited to the UK and the methods presented here can be replicated in other settings where local data are unavailable, to support planning and prioritisation of food aid resources.

The short and long-term health implications of food insecurity are not well assessed in the UK, in part because systematic measuring of food insecurity prevalence in the population only started in 2019 as part of the Family Resource Survey (FRS). This followed substantial campaigns by the third sector and academics (14, 15). Results from the first FRS dataset indicated that in 2019/20, 5 million people (8%) were living in food insecure households (16). Earlier data on food insecurity were captured in the Food and You Survey in 2004 and 2016 (17). In this study of the two datasets collected 12 years apart, unemployment, low income and disability were all associated with severe food insecurity while younger age, non-white ethnicity and low educational attainment were also associated with food insecurity. Surveys conducted by the Food Foundation (18) during the 2020 Covid-19 pandemic, and as part of the longitudinal Understanding Society (19) study, provided further indications of who experiences food insecurity in the UK, noting associated health outcomes including mental wellbeing. A UK-level assessment of food insecurity using a slightly different measure identified similar sociodemographic risk factors, including people on lower incomes and those renting their homes (20). Despite these recent efforts to collect more data on the extent of food insecurity in nations of the UK, a lack of consistently collected subnational data remains.

### Measures of risk

In the absence of known prevalence of a health outcome or social phenomenon the estimated risk to the population to experience the outcome is often predicted; in 2020 this was demonstrated for diabetes in small areas of England (21). Within the UK, area-based measures are used to aid prioritisation of resources. The most commonly applied sociodemographic measure is the Index of Multiple Deprivation (22) with the most recent release in 2019. This is a wide-ranging index with seven domains/groupings of variables (including income, crime, education, health) to reflect relative social and material deprivation at the scale of Lower Super Output Areas (LSOAs) across England. These small areas have populations of about 1500 and there are 32,844 in England. Similar indices exist for Scotland (Scottish Index of Multiple Deprivation) and Wales (Welsh Index of Deprivation) for small areas (23, 24). Often these indices of deprivation are used as shorthand for poverty or (inaccurately) as proxy indicators of individual socioeconomic status for residents (ref). They provide a useful tool for local government organisations who are tasked with planning and distribution of resources for their local populations. The IMD offers a good general indicator of relative deprivation with supplemental indices focused on older people and children, however, they include 39 indicators across the 2019 Indices of Deprivation (25). When poor health is associated with social and material deprivation, these deprivation measures offer a reasonable proxy for risk of ill health in local populations, and are used in individual level risk scores for poor health (see QDScore (26)). For more specific purposes, the IMD may be overly complex when the researcher is interested in one outcome, such as disordered eating, which may not be easily predicted by a composite measure (or one of the constituent domains). An additional challenge is the risk of multicollinearity between variables in a regression analysis, for example using the overall IMD values as well as distance to food stores when exploring predictors of excess weight. There is scope for bespoke area-based measures of population characteristics, health and risk to support local governments with planning for health and social care, and for third sector organisations for locating interventions such as food aid.

There have been efforts in recent years to develop targeted area-based indicators of health risk and deprivation, to better reflect the experiences of specific populations or to focus on selected concerns such as diabetes. In response to an awareness that the ‘standard’ English IMD measures did not reflect rural populations, Burke and Jones developed an index of rural deprivation (27) which was welcomed by local governments in the East of England as an improved assessment of their populations’ challenges. This measure included more variables on the role of service access, which comprises less than 10% of the standard IMD (25). Two other measures explored the environment rather than population characteristics. The first, devised in 2010, created a measure of environmental deprivation related to health for the UK (28) and was focused on assessing the impact on health outcomes in populations from the local environment. This measure included hypothesised health promoting and health damaging characteristics of the natural environment including air pollution, UV radiation and green spaces. The resulting index, Multiple Environmental Deprivation Index (MEDIx) may be used similarly to the IMD. This measure is shown to be associated with mortality across the UK, demonstrating the relevance of measures which are focussed on specific aspects of localities(29).

Further development in this area of research is a UK-based classification of local environments to understand the potential impacts of place on population health, the Access to Healthy Assets and Hazards (AHAH) index. In AHAH, LSOAs are categorised based on access to a range of amenities and natural environments which are described as potentially beneficial or detrimental to health (30). AHAH incorporates variables across domains of access to health services, retail outlets and environmental quality. A substantial benefit of AHAH is that the data are all freely available to researchers. AHAH was found to be associated with mental wellbeing, though not associated with self-rated health or limited long term illness (30).

In addition to the earlier version of local area food insecurity risk developed in 2018 for England, two further measures of food insecurity risk were developed which represent estimates of food insecurity in the UK. The first makes use of machine learning approaches to model food insecurity based on users from the Olio food sharing app (31). This measure took the demographic profiles of users who asked for food and applied machine learning to a sample of 421 app users for training and testing algorithms, which formed the basis of further models. Area data in this model included proximity to food stores, bus stops and food banks. This risk measure is created at LSOA level, at the time of writing the data are not freely accessible. A challenge with these data is the reliance on the small sample of app users which may over-represent a unique population who are unlikely to be active outside of urban areas where food sharing is more accessible or feasible.

A local authority level model was devised from data collected by the Food Foundation (n=4231 survey respondents) in January 2021 which applied small-area estimation methods to estimate differing levels of food insecurity and hunger (32). This application of estimation approaches is novel in modelling food insecurity, with the benefit of confidence intervals around the estimates to reflect uncertainty. Modelled data are available to download or in map format, enhancing accessibility for colleagues working in the national government or policy. These data represent the prevalence of people estimated to be hungry, to struggle or who worry about food. Like the AHAH data and the previous food insecurity risk measure, the availability of these data is a strength of the research. The main limitation is the scale of the estimates at local authority level (343 are present in England, in contrast to 32,844 LSOAs), which are beneficial for national or regional level prioritisation, however, finer scale estimates are needed within local government.

### Aim

The aim of this study is to develop an updated measure of food insecurity risk for small areas in England (11) using data which are open access and enabling users to differentiate risk between broad area types (urban and rural). The methodological approach is outlined and the patterns in the resulting measure(s) described. The level of agreement between this risk measure and related measures or outcomes at LSOA and Middle Super Output Area (MSOA) (typical population of 7000) levels is included for validation.

## Materials and Methods

The process of developing a new version of food insecurity risk measures (hereafter the Complex Index) was informed through a scoping/literature review and semi-structured interviews with stakeholders in Wessex (Hampshire, Southampton, Portsmouth, Dorset, Isle of Wight), England. These stakeholders included people working in food banks and local government, who were able to reflect on the demographic characteristics of people seeking food aid. Although not all people who identify as food insecure access food aid formally (33), these interviews held between December 2020 and February 2021 were intended to identify prevailing characteristics of people experiencing food insecurity and seeking help with accessing food due to economic constraints. Interviewees were identified through contacting local government teams across Wessex for introductions to appropriate members of staff and local food bank coordinators. The research was granted ethical approval by the University of xx (blinded for peer review) Ethics and Research Governance Online (ref 55822.A1).

We undertook a scoping exercise to identify relevant data to consider for inclusion in this risk measure. The previous food insecurity risk measure was developed from literature review of studies that explored self-reported food insecurity, including Kneafsey and colleagues (34). The original measure included two domains of data: demographic characteristics of people at higher risk and the population claiming welfare benefits, all expressed as a percentage of the population living within MSOAs. Higher risk included people aged 65 years and over who lived alone or those under the age of 65 who are on low incomes and have dependent children, as a percentage of the relevant population. An interim update in 2020 added single adult households under the age of 65 on a low income. More recent literature identified further sociodemographic indicators of food insecurity risk including poor mental health and disability (17, 35).

Here, as part of a wider project, we updated the earlier measure (referred to now as the Simple Index) with newer data (Department for Work and Pensions benefit claimant counts from May 2020) to produce updated local maps of estimated risk. We conducted semi-structured interviews (n=11) with local stakeholders across Wessex and email discussions with regional and national level policy colleagues and contacts in other local authorities. Interviews were conducted using Microsoft Teams between December 2020 and February 2021 by two researchers (GG and DS), transcribed and key recurring themes were identified from the transcripts. As part of the interviews and wider email contacts, participants were asked to look at an MSOA level map of the Simple Index and comment on whether the risk pattern in their local area reflected what they observed in their roles as food bank coordinators, local government leads for food poverty or welfare benefits. Interviewees were also asked for their observations about clients accessing services for food aid to gain insight into the demographic characteristics of households accessing support. We used these qualitative interviews and the ongoing discussions held with colleagues working in public health or policy as the basis for updating the risk measures. The variable selection was informed by these interviews and discussion as well as newer literature available in the topic area (17, 19).

Interviewees reported the most frequent household structure, size and employment status of people accessing or seeking support for food insecurity. Common themes included access to employment, transportation, digital exclusion, cost of food, cost of housing, energy bills. Further recurring themes were poor health including mental health, disability, lack of skills needed for better paid employment.

We first updated the Simple Index with the most recent data, at the lower LSOA scale (Table 1). This consisted of two domains. First, the percentage of individuals in an LSOA receiving any welfare benefits appropriate for their age group, sourced from the Department for Work and Pensions datasets (see https://stat-xplore.dwp.gov.uk/webapi/jsf/login.xhtml). These datasets avoid any double-counting of benefit recipients by taking account of all possible benefit combinations. The second domain was household composition which included the percentage of lone pensioners (here, people aged 65 and over) or in the under 65 population, low-income households with lone individuals or with dependent children. Low income was defined by using Census 2011 data on the National Statistics Socio-economic Classification (NS-SEC) where the household representative person was employed in the three categories of semi-routine occupations, routine occupations or never worked and long-term unemployed (Table QS608EW).

**Table 1.**
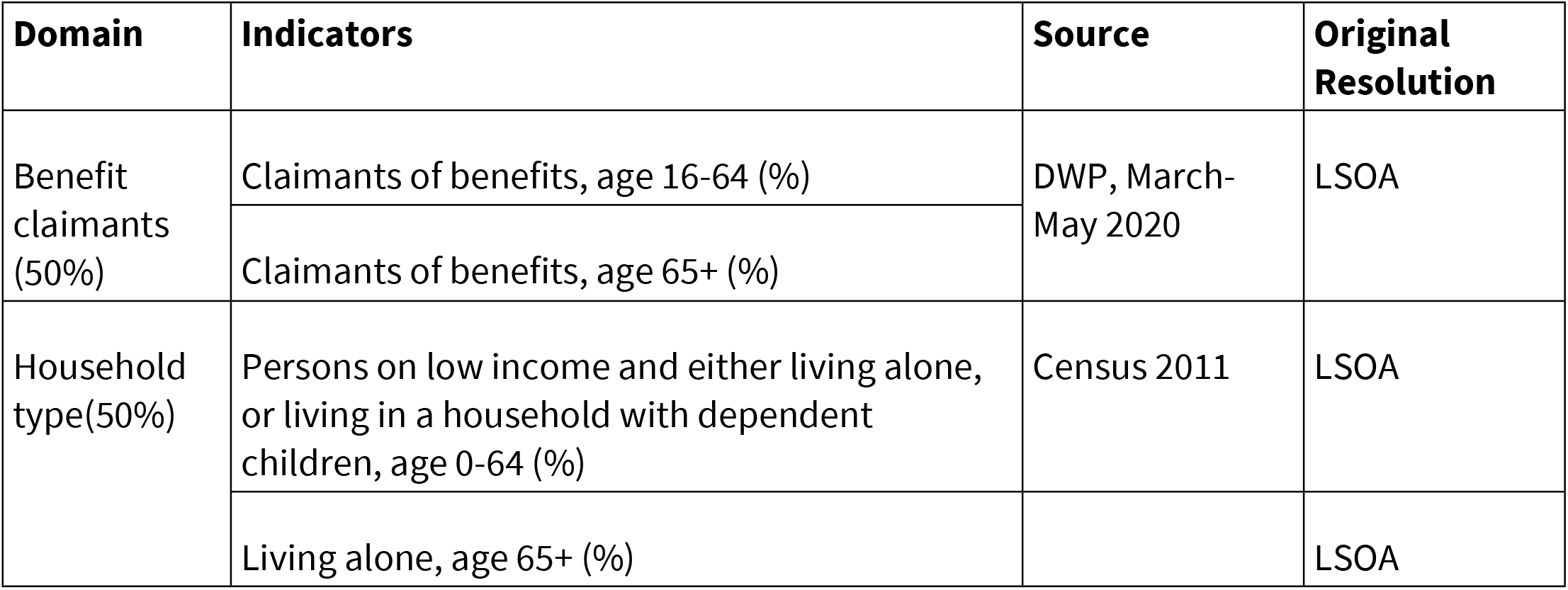
Simple Index Domains and Indicators.

| Domain | Indicators | Source | Original Resolution |
| --- | --- | --- | --- |
| Benefit claimants (50%) | Claimants of benefits, age 16-64 (%) | DWP, March-May 2020 | LSOA |
|  | Claimants of benefits, age 65+ (%) |  |  |
| Household type(50%) | Persons on low income and either living alone, or living in a household with dependent children, age 0-64 (%) | Census 2011 | LSOA |
|  | Living alone, age 65+ (%) |  | LSOA |

A Complex Index was developed using further indicators identified through the interviews and literature review (Table 2). We considered two categories of additional factors: Compositional, focusing on population characteristics, and Structural, which describe attributes of the LSOAs such as access to services.

**Table 2.** Complex Index Domains and Indicators.

| Domain | Indicators | Source | Original Resolution |
| --- | --- | --- | --- |
| Compositional (individual characteristics) (50%) | Claimants of benefits, age 16+ (%) | DWP May 2020 Working age and Pension Age claimant groups | LSOA |
|  | Persons on low income and either living alone, or living in a household with dependent children, all ages (%) | Census 2011 | LSOA |
|  | Persons with no educational qualifications, age 16+ (%) | Census 2011 | LSOA |
|  | Mental ill health, composite | IMD 2019 | LSOA |
| Structural (area characteristics) (50%) | Minutes to nearest employment centre (size 100+ jobs) by public transport (bus, train), age 16-74 | Department for Transport 2017 | LSOA |
|  | Median download speed Mbit/s by connections in an area | Ofcom Fixed performance data 2020 | OA |
|  | Bus stops per km<sup>2</sup> using LSOA area size from the ONS | National public transport access node (NaPTAN) 2020 | Coordinates |
|  | Distance (Euclidean km) to medium and large grocery stores (1,400m<sup>2</sup>+) | Geolytix Retail Points 2021 | Coordinates |

The Compositional Domain includes the Simple Index composed of benefit claimants and household demographics. This is extended with the inclusion of the percentage of individuals without any educational qualifications (from the Census 2011 table LC5102EW) and the LSOA-level mental health score derived from the Mood and Anxiety component of the IMD 2019 health domain(25).

The Structural Domain includes geographical barriers that directly or indirectly impact food insecurity risk. This includes restrictions produced by public transport reliance with indicators for distance to larger food stores, travel time to employment centres and area bus stop density. Indirect structural impacts on economic status are considered through local internet speeds (digital exclusion).

The inclusion of these indicators was based on the scientific literature and stakeholder interviews as described above. The overall aim was to produce a composite index of risk to enable ranking of neighbourhoods using multiple data sources that are open access, can be quickly updated and easily interpreted.

### Data Collection and Preparation

Data for the indicators were collected from open sources at the area level closest to LSOA for England (n 32,844 areas) and the most recent data published.

For the Simple Index, which follows the example from (Author, 2018) benefit claimant data were updated from the Department of Work and Pensions (DWP) for 2020 using the ONS mid-year population estimate for 2019 (all ages) as its denominator, as some benefits may be claimed on behalf of children under age 16. The data were extracted for Benefit Combinations, Working Age Caseload and State Pension Age Caseload. Benefit data are for the previous three months, ending May 2020. Although this time period captures the beginning of the 2020 Covid-19 Pandemic, further analyses with datasets over several years showed little change in the ranking of LSOAs based on the prevalence of benefit claimants. These data were the most up to date at the time the indicator was developed. Household composition data originated from the Census 2011, using two tables to calculate a joint probability of people under age 65 to be in a low-income household (from table QS608EW) either as a single adult or with dependent children (from table DC1109EW). This is summarised in Table **1**.

For the Complex Index, multiple sources of data were consulted. For the Compositional Domain this included the two indicators from the Simple index, the Census 2011 table for highest educational qualification (Table LC5102EW) and the IMD 2019 data for LSOA level Mood and Anxiety disorders indicator. This IMD composite indicator includes data from hospital episode statistics, prescription data and suicide mortality rates to provide a broad measure of mental ill health in local areas (25). We explored options to include BAME populations from the 2011 Census as one indicator. We did not use it in this Domain because in earlier assessments it did not correlate well with our validation data.

For the Structural Domain, data were collected with a focus on access via public transport (bus, train, walking) to highlight deprivation produced from not owning a car. This includes travel time to job centres of more than 100 employees for the working age population, originally derived from the ONS Business Register Employment Survey (36). Median download speed was derived from annual statistics provided by Ofcom (37). Although this measure includes business connections, download speed was chosen over a measure of residential service availability as the majority of homes were above the Universal Service Obligation (>10Mbit/s download speed) and users may not be able to pay for the fastest internet speeds available in their area (37).

A count of bus stops per square km was created using point data (38) to account for the variation in area size of LSOAs, where the locations of bus stops were projected into LSOA boundaries. The Euclidean distance to medium and large grocery stores was derived from open data provided by Geolytix, a private data company (39). The use of larger stores was based on previous research showing that smaller format stores provide limited choice or quality often at inflated prices (40). Euclidean distance rather than network distance was used due to computational power. This is not a limitation as the two measures (Euclidean and network distance) are strongly correlated in urban settings (41). The indicators of the Complex index are summarised in Table 2.

Although a housing affordability indicator was developed using median house price paid at the LSOA level and average income data at the MSOA level (42) this indicator was not used as it was found to skew the data towards unaffordability in high income areas and affordable housing available in low-income areas. The IMD housing affordability indicator was not used as it represents only the under age 40 population (25) and other housing statistics such as rental prices were not available at LSOA level, or had high levels of missing data.

### Data Processing

When all indicator data was prepared at the LSOA level, each indicator’s data was transformed to a normal distribution using the Rankit method, a rank-based inverse normal transformation (43). This was to prepare data from different sources to be combined into a single domain. All indicators were normalised in the same direction so that the smallest score value represents the theorised most risk of food insecurity for that indicator. For example, the LSOA with the lowest count of bus stops and longest distance to stores would each represent the smallest score for those indicators. For the mental health indicator, the Mood and Anxiety score from the IMD (where deprivation is indicated by the highest score in this index) was normalised to match the direction of other scores with the smallest score indicating the highest risk of mental ill health.

Relevant indicator scores were summed for each LSOA to produce a domain score. Equal weighting of scores was chosen due to a lack of evidence suggesting a greater emphasis on any particular measure (44). Domain scores were prepared for combining into an index using an exponential transformation, as used in the creation of the IMD and elsewhere (43). This was applied to reduce any cancellation effects whereby low risk scores in one domain may ‘cancel’ high risk scores in the second domain. This transformation also emphasises LSOAs at higher risk of food insecurity which facilities identification. The exponential transformation of a domain score (X) is calculated as follows:

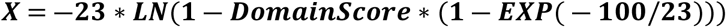

Where ‘LN’ denotes the natural logarithm and ‘EXP’ the exponential or antilog transformation. 23 is the scaling constant to minimise cancellation.

The exponentially transformed domain scores are then added together to produce a composite index score using equal weighting (Complex Index). This score is then ranked for each LSOA where 1 represents the lowest score and therefore the population most at risk of food insecurity. This ranking provides a more interpretable scale of the risk of food insecurity for a multi-dimensional index than its score. For areas with identical risk scores, the lowest applicable rank is applied to both areas. Data were processed using SPSS Ver. 26.

#### Validation

The Simple Index, Complex Index and its domains were compared with the ranks of other indicators and outcomes known to be associated with food insecurity to assess validity (44) (6) (Table 3). This includes the rank of IMD 2019 already at LSOA level as well as two further variables associated with household food insecurity, child obesity prevalence and free school meal uptake.

**Table 3.**
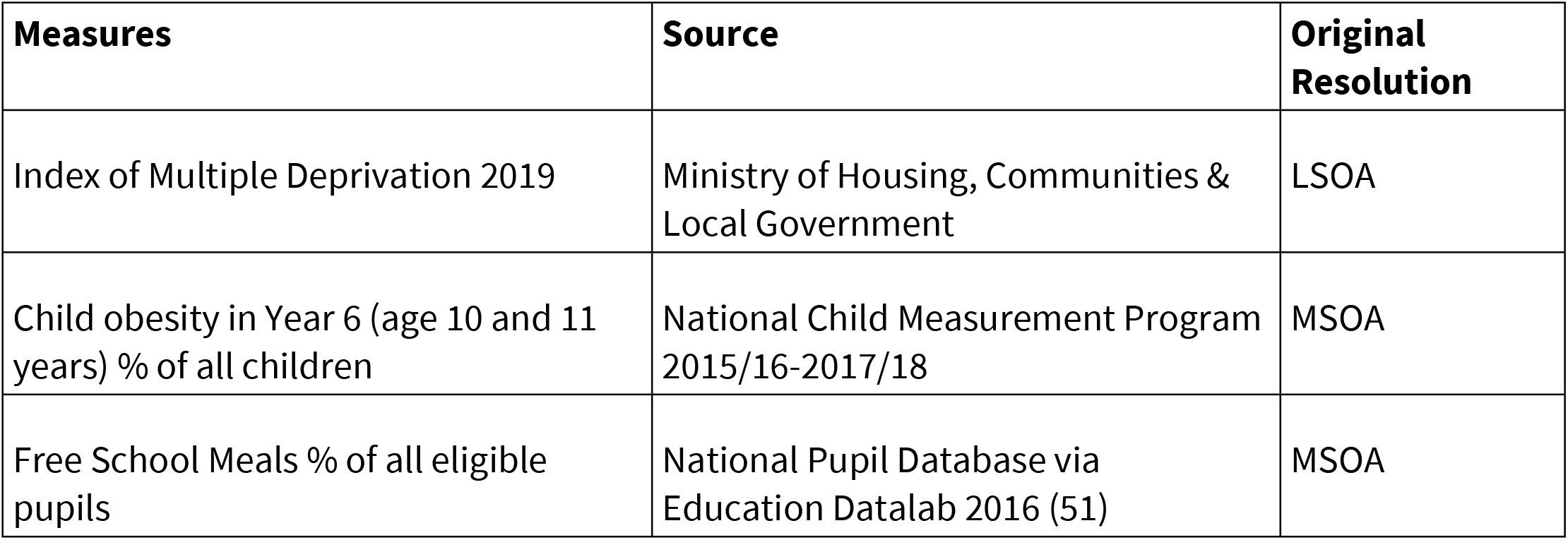
Validation Measures.

The previous index was validated using data on child weight and compared to the 2015 IMD score, as there is a known relationship between material and social deprivation as measured in the IMD and food insecurity (45). Childhood obesity is associated with food insecurity (46-48), due to the quality of food available in food insecure households. Access to free school meals in England is based on household income and benefits claimed, including Income Support or Child Tax Credit. This also includes Universal Credit, but household income must be below £7,400 a year if applying after April 2018 (See https://www.gov.uk/apply-free-school-meals). Recent research reported an association between receiving FSM and use of food banks (19), with the use of food banks often used as an indicator of household food insecurity. However, due to the relatively low uptake of food banks in households otherwise classified as food insecure in Canada (33, 49), food bank use is not used as a proxy measure for food insecurity in this study.

Measures of the percentage of child obesity (year 6, average value across three years 2015-18 from National Child Measurement Programme) and percentage of pupils eligible for free school meals (2016) were only available at MSOA level as shown in Table 3. For comparison with these two indicators, our index was aggregated to MSOA level. This was developed through the summation of exponentially transformed domain scores at LSOA level to their relevant MSOA, as indicated in area lookup tables provided by the ONS. This MSOA level index and validation indicators were ranked with 1 being the MSOA with the least desirable outcomes for any variable (highest risk of food insecurity, highest prevalence of child obesity) to enable comparison.

Ranks of the Complex Index and its two domains (Compositional and Structural) were compared to ranks of the validation variables. Analysis used Spearman Rank correlation for non-parametric scales. Deciles of ranks were also developed for comparison categorically using Chi-square tests (χ^2^). One of the concerns raised by previous researchers is the ability of indices to reflect both urban and rural populations(27). To assess the fit of our measures we repeated the correlation and chi-squared analysis for all LSOAs, those classified as urban, and those classified as rural as defined by the ONS 2011 (50). Intraclass correlation coefficient (ICC) was used to measure agreement between the area ranking of our index and rankings of selected measures for validation. Measures of absolute agreement were estimated using two-way mixed effects models, assuming different fixed observers. The ranks of the individual variables comprising the Compositional and Structural Domains were correlated against their relevant domain rank to ensure variables were correlated in the expected direction.

## Results

Following the development of the indices and domains (Simple, Complex Indices; Composition and Structural domains of the Complex Index), we assessed how well these outputs aligned with the validation data. With favourable outcomes, we have identified a recommended approach to identifying food insecurity risk in most areas of England and have provided suggestions on mapping the two domains of Compositional and Structural alongside each other in rural areas. The final ranks of LSOAs for the indices and domains are available online at (blinded for peer review).

Our original index published in 2018 was found to correlate well with the Index of Multiple Deprivation 2015 (IMD), as does this updated Simple Index at a finer resolution for the IMD 2019 (r_s_=0.872, ICC=0.932). This is supported by a significant χ2 value (51,961), when considering deciles of risk observed in Table 4. The Simple Index also correlated well with the 2016 prevalence of free school meal eligible pupils (r_s_=0.812) and year 6 (age 10-11) obesity prevalence (r_s_=0.730), when ranked by small areas. This confirms informal observations from local authority teams across the country that our simple measure accurately reflects the areas where more of the population is experiencing food insecurity.

**Table 4.**
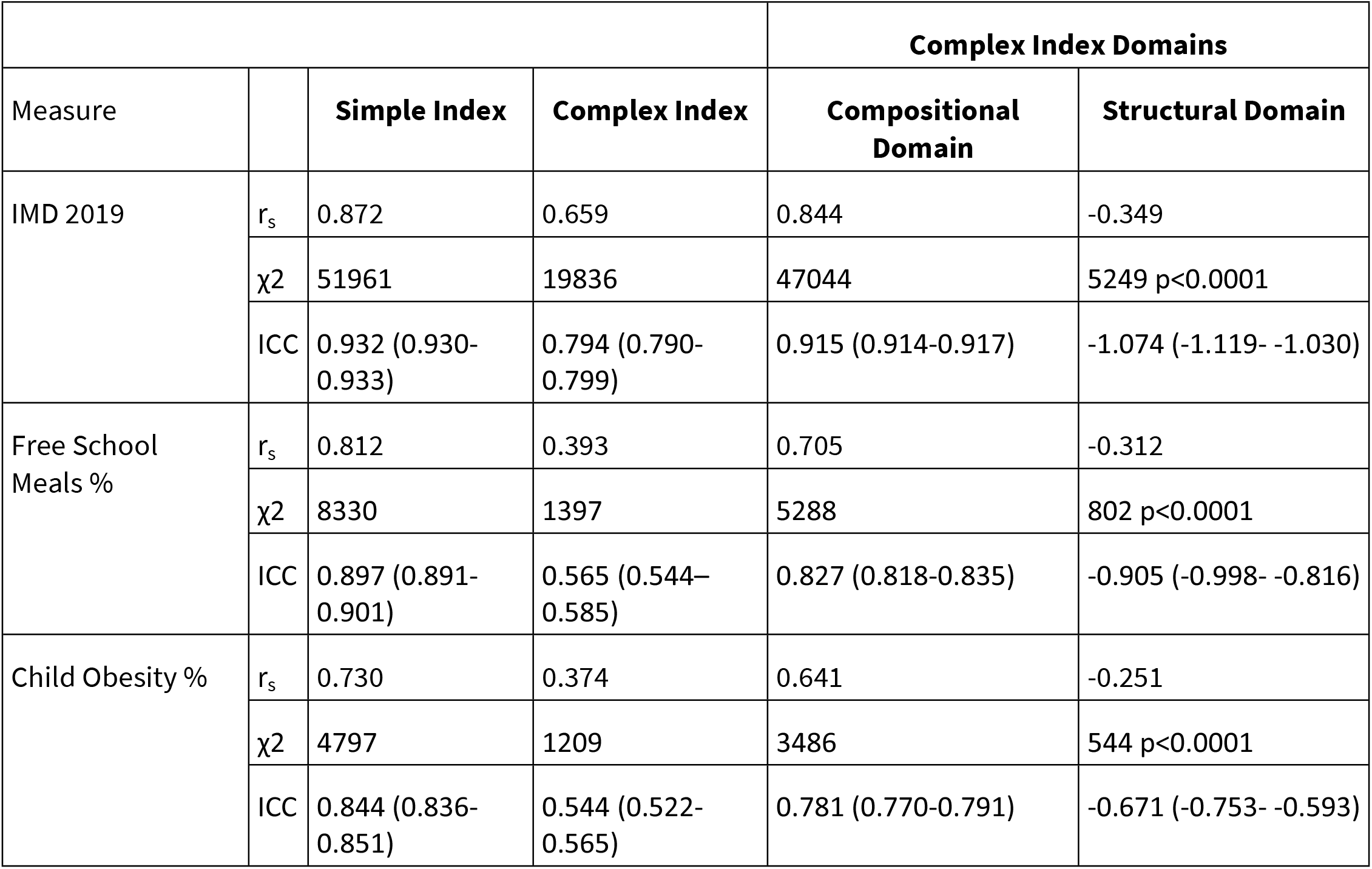
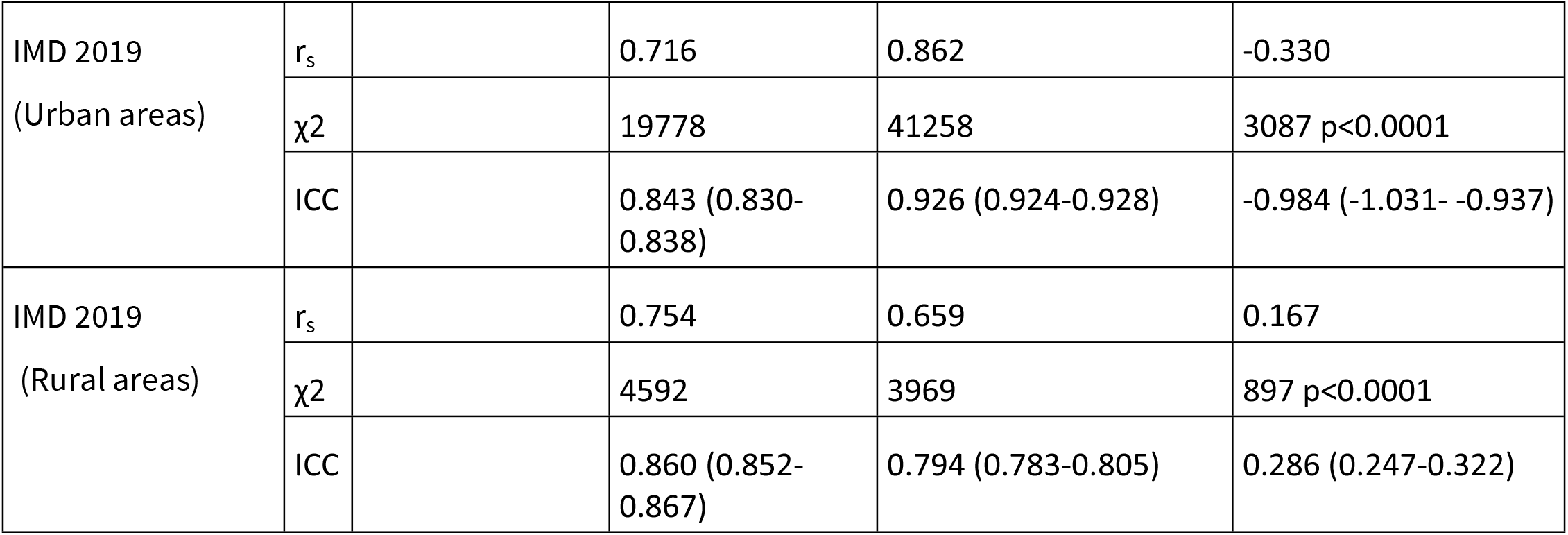
Validation Results. r_s:_ Spearman Rank, χ2: Chi-square ICC: Intra-class correlation

As noted above, the Complex Index and its two domains were assessed for correlation with all LSOAs and then for LSOAs classified as urban and rural areas separately. The Complex Index correlated moderately with the IMD 2019 (r_s_=0.659), with a weaker χ2 value (19,836) and ICC (0.794) when compared to the Simple Index. The Complex Index also correlated weakly with the percentage of free school meal eligible pupils (r_s_=0.393) and year 6 obesity rates (r_s_=0.374). When restricted to areas classified as rural, the correlation of the Complex Index with the IMD 2019 improves (r_s_=0.754, ICC=0.860) with urban areas slightly less well correlated (r_s_=0.716, ICC=0.843). However, urban LSOAs χ2 values are similar to the Complex Index (19,778) with rural area χ2 values much smaller but still significant (4,592).

Considering the separate domains, the Compositional Domain for all areas (r_s_=0.844, ICC=0.915), and for urban areas alone (r_s_=0.862, ICC=0.926), correlates very well with the IMD 2019 and is supported by χ2 values close to that found for the Simple Index (domain χ2 = 47,044) (Table 4). For rural areas alone, the Compositional Domain is more moderately correlated (r_s_= 0.659) with a small but significant χ2 value (3,969). The Compositional Domain demonstrates a stronger correlation with free school meals (r_s_=0.705) and obesity rates (r_s_=0.641) compared to the Complex Index.

The Structural Domain alone is negatively correlated with validation measures, except the IMD 2019 in rural areas (r_s_=0.167, ICC=0.286), and has weak correlation and small, but still significant, χ2 values with all validation measures.

For the Simple Index, 26% of areas in the top decile of risk were in the North West with 97% of these areas classified as urban. For the Complex Index, neighbourhoods in the top 10% most at risk (n=3284) were found across all regions of England with the highest percentage in the North West (25.5%). In this top decile, 74.4% were in urban areas. For the top decile of risk in the Compositional Domain, 32.5% of the LSOAs were located in the North West region with 96% in urban areas. For the top decile of the rural only index, highest risk LSOAs were more often in the East of England.

As part of the preparation of this updated index we shared the resulting maps with our stakeholders and asked for feedback about which aligned best with their local knowledge. Across all areas of Wessex and Lancaster, the most accurate maps were identified as the Compositional Domain. We have mapped these results below in Figure 1.

**Figure 1:**
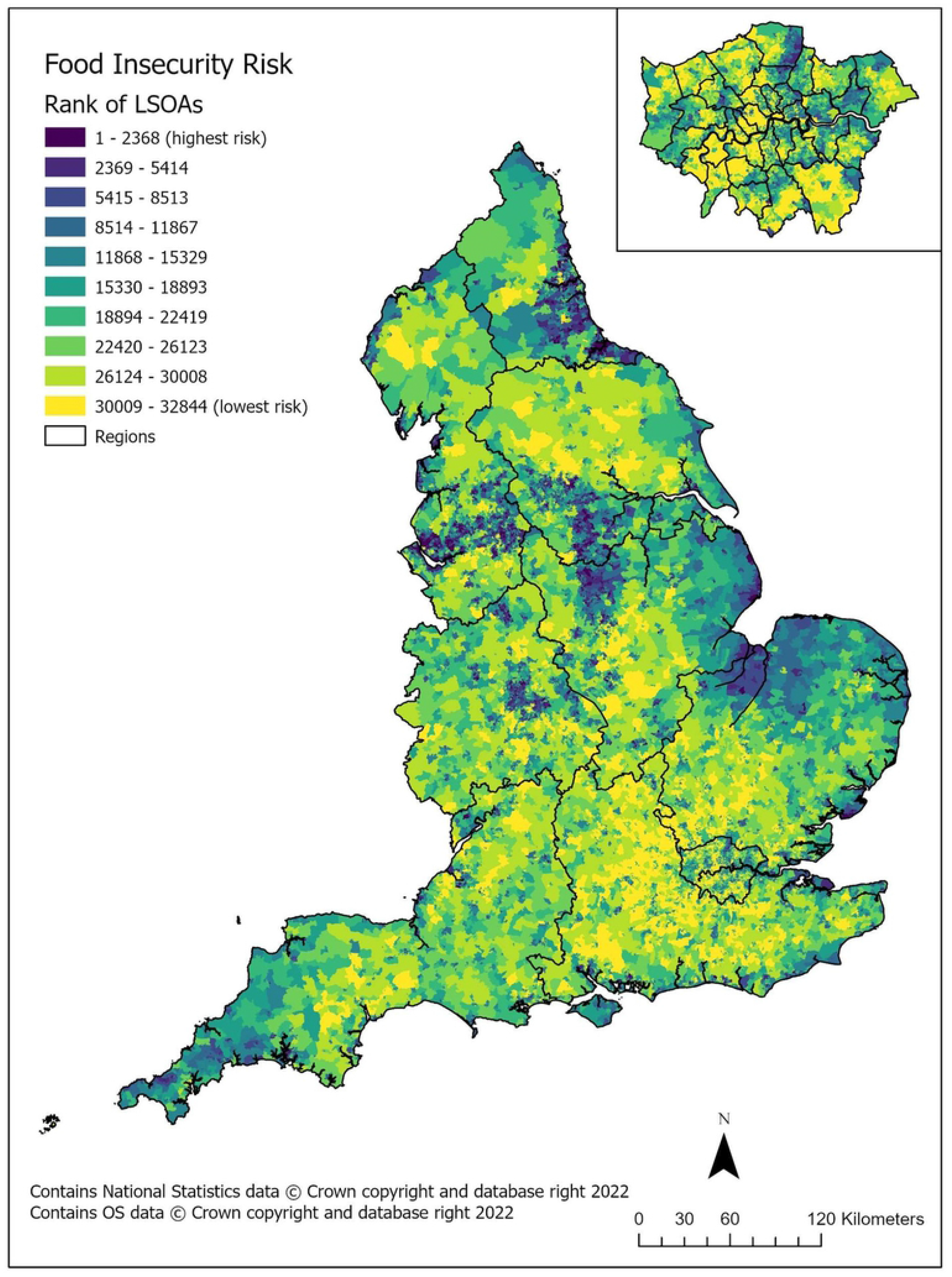
Food insecurity risk (compositional domain) in England by LSOA, deciles, including region boundaries. Inset map of London.

Our recommendation is for most teams working in local authorities, or in the third sector, to use the Compositional Domain as the primary measure when exploring patterns of food insecurity risk in their local populations. Given the impact on food security that access to amenities and services can have in rural locations (52), we suggest that a map of the Structural Domain is included alongside the Compositional Domain in these areas. The Structural domain as part of the Complex measure emphasises structural factors of public transport, access to stores and faster internet connections and employment. These will present a larger challenge for more rural locations, however, these are very real challenges for people living in rural areas.

## Discussion

Updated indices of household food insecurity risk were collaboratively developed to support charities, health and public sector (especially local government) organisations. The development of these indices were informed by a scoping review, similar to AHAH (30), and in discussion with end users, like the rural IMD update (27). After consultation with stakeholders, a series of indicators were assessed for inclusion in an updated measure. The main criteria were relevance to published research on social and demographic predictors of household food insecurity and for the data to be open access to facilitate regular updates of the new indices. Some flexibility to reflect risk in urban and rural settings was desired, as often multidimensional deprivation indices are more focused on the urban experience (27). The full list of possible variables was reduced on the basis of completeness of datasets, availability of LSOA-level data and the impact on modelled correlation, ICC and chi-squared analyses.

The resulting options have been shared with our stakeholders and wider network, with enthusiasm for the new resources that capture more aspects of the circumstances which contribute to household food insecurity, such as mental health. This is confirmed in our approach to validation and is consistent with previous research into the contributing factors of food insecurity which identifies poorer mental health as a risk for household food insecurity (53-57). Educational attainment was the other new variable added to our recommended Domain (Compositional), which was a common theme from our stakeholder interviews and confirmed in the literature as well (17). Here it represents limited options for higher earnings, improved employment as we selected the prevalence of the population with no educational qualifications. The original components of the 2018 index remained relevant, benefit claimants being consistently more likely to access food aid, at times due to challenges with benefit claims (35, 57-59).

Further considerations that arose from the interviews were the geographic inequalities/locational disadvantage that contributed to food insecurity, particularly in more rural settings. These included the cost of housing, access to transportation and access to appropriate employment. As part of this, the ‘digital divide’ which describes limited access to the internet was reported. We compiled data to reflect these structural factors and were able to develop a domain that represents these challenges in accessing affordable food, public transportation, online access and employment opportunities. Although not included in the measure recommended for most areas (Compositional Domain) this Structural Domain does capture similar data as the updated rural IMD score developed in Norfolk (27) and is aligned with the AHAH index that reflects local physical environments and amenities (30, 43). The inclusion of the AHAH measure in national resources (PHE Fingertips provided by the Office for Health Improvement and Disparities [OHID]) demonstrates the relevance of using built environment data for planning and policy, where there is concern for public health.

Selecting the most appropriate measure was informed by the validation process and consultation. The Complex Index reveals that most areas are not in the highest decile of risk for both Compositional and Structural domains. Rural areas may be at risk structurally, but residents may be able to offset the issues caused by a longer distance to services where there is *reduced* household economic risk. For example, if households have access to a car, then public transport is likely to be less essential. This reflects an issue of locational disadvantage for residents of areas where most households are better off financially, as there are fewer resources to offer support for those who do need it. Our suggestion is for the Composition Domain to be applied in most areas, as it adds further data from the Simple Index and was identified as fitting well with local knowledge when shared with wider audiences across England.

A challenge with any area-based measure is that areas and their populations are classified in a way that may miss individual experiences. We are aware that rural areas may have a high prevalence of wealthier households, however, there will be pockets of deprivation that are masked when using composite risk scores. In rural areas this may pose an even larger problem for households facing difficulties both due to the lack of access to support noted above and the inability to anonymously seek assistance. Households in urban settings may have better access due to a higher concentration of food aid or welfare advice resources, also better links for public transport for households without cars.

### Strengths and Limitations

Using several data sources in the Complex Index reduces the bias that may be produced from considering only one dimension of food insecurity risk. The variables included are sourced predominantly from open access validated governmental statistics and data. We assessed a wide range of variables for suitability as outlined above and drew upon a network of contacts to ensure the resulting indices were applicable in densely populated urban settings as well as sparse rural areas, and diverse populations. Feedback was gained from a wider audience outside of Wessex and the South East at seminars and presentations to audiences in London and online between December 2019 and April 2021. This work reached colleagues in all regions of England and international audiences, facilitating discussion of the indices for applicability across the country.

Data are available to download or map using a website (blinded for peer review). This was co-developed with third sector and local government partners to enable access to the data and resources for visualisation where mapping software and expertise is not easily available. This will allow uptake by more organisations, supporting the further inclusion of these risk indcies in annual reports such as Joint Strategic Needs Assessments (JSNAs) or funding applications for food aid interventions. Correspondence with users of the indices suggest these are key applications of the data (private correspondence, January 2021).

As with any small area measure, there are limitations. DWP data applies statistical disclosure which may impact the accuracy of percentage calculations for benefit claimants. However, this is partially addressed as we do not use raw data in our development of indices. Three variables (qualifications, household structure, low-income households,) were informed by the 2011 Census that will benefit from updates from the more recent census as it becomes available; the risk indices are updated annually in September. The 2019 IMD score includes one data point (mental health) also included in this measure, so there is minimal risk of collinearity in the validation process. However, the mental health indicator in the IMD score comprises only 2.3% of the overall score for an LSOA. Benefits claimant data included in the 2019 IMD score are from a different time period and for specific benefit types only (25).

The ranking of an LSOA is not an absolute measure of household food insecurity but can be used to compare against the ranking of other LSOAs, particularly useful for illustrating areas at relatively higher risk. Although the indices are measured at the local neighbourhood level, the LSOA rank may not reflect the situation of every local resident.

## Conclusion

The work presented here provides an important update and improvement on an earlier measure of food insecurity risk using a finer local scale and greater flexibility in the data used to assess risk in local areas. Initial discussions with stakeholders in local and national government (as part of the scoping for this research) highlighted the need for a measure at the same scale as the IMD, to provide the specificity required in local areas. These risk indices can be adapted for other settings, including the other nations of the UK or countries including Australia and New Zealand. Data such as number of benefits claimants in an area are updated regularly, and as a result the indices are updated annually.

Household food insecurity is a problem facing many households in England. Longer term solutions to the situation are needed, including supporting households to maximise their incomes and addressing structural barriers to food security. In the shorter term there is a proliferation of food aid and assistance offered across local areas; this research supports decision-makers to target assistance where it is likely to be most needed.

## Data Availability

The data underlying the results presented in the study are available from https://mylocalmap.org.uk/iaahealth/

## Acknowledgements

We would like to thank all participants in this study for their time and engagement with the interviews and further conversations.

